# The Vaccination Threshold for SARS-CoV-2 Depends on the Indoor Setting and Room Ventilation

**DOI:** 10.1101/2021.07.28.21261300

**Authors:** A. Mikszewski, L. Stabile, G. Buonanno, L. Morawska

## Abstract

**Background:** Effective vaccines are now available for SARS-CoV-2 in the second year of the COVID-19 pandemic, but there remains significant uncertainty surrounding the necessary vaccination rate to safely lift occupancy controls in public buildings and return to pre-pandemic norms. The aim of this paper is to estimate setting-specific vaccination thresholds for SARS-CoV-2 to prevent sustained community transmission using classical principles of airborne contagion modeling. We calculated the airborne infection risk in three settings, a classroom, prison cell block, and restaurant, at typical ventilation rates, and then the expected number of infections resulting from this risk at varying levels of occupant susceptibility to infection.

**Results:** We estimate the vaccination threshold for control of SARS-CoV-2 to range from a low of 40% for a mechanically ventilation classroom to a high of 85% for a naturally ventilated restaurant.

**Conclusions:** If vaccination rates are limited to a theoretical minimum of approximately two-thirds of the population, enhanced ventilation above minimum standards for acceptable air quality is needed to reduce the frequency and severity of SARS-CoV-2 superspreading events in high-risk indoor environments.

## Introduction

Control of infectious disease is achieved when the average case does not beget another, and transmission becomes sporadic in nature. For airborne contagion in shared indoor atmospheres, Wells [1] established that the rate of transmission is inversely proportional to the ventilation rate per susceptible occupant. It then follows that to control airborne contagion we can either increase ventilation, or its equivalent through air filtration or disinfection, or decrease the number of susceptible occupants through vaccination [2]. During the COVID-19 pandemic, lockdowns and occupancy controls have been widely applied to reduce transmission of SARS-CoV-2. These are blunt but effective methods of increasing the ventilation rate per susceptible occupant of indoor spaces. As SARS-CoV-2 vaccines become available to the public in 2021, the question becomes: at what point is the number of susceptibles in public spaces low enough so that occupancy limitations are no longer necessary to control the spread of the virus?

To address this question, we must consider the primary settings of SARS-CoV-2 transmission. As with other agents of airborne contagion such as *Mycobacterium tuberculosis*, SARS-CoV-2 thrives in congregate living and working spaces with shared air, such as prisons, schools, restaurants, abattoirs, and care homes. The COVID-19 pandemic is also fueled by superspreading events in crowded indoor environments where people vocalize and cannot reliably wear masks. For example, Chang et al. [3] modeled full-service restaurants to produce by far the largest increase in infections upon reopening after lockdown of any non-residential location that people visit. Estimates of necessary vaccination rates for these high-risk community settings should be protective in other microenvironments, and therefore approximate a vaccination threshold to control SARS-CoV-2 such that the average case fails to beget another.

The aim of this paper is to estimate setting-specific vaccination thresholds for SARS-CoV-2 using classical principles of airborne contagion modeling. We included modeling scenarios for a prison cell block and a full-service restaurant, two settings known to be high risk for SARS-CoV-2 transmission. To compare the vaccination threshold for SARS-CoV-2 to historical estimates for measles virus, we also included a classroom scenario in our analysis. A secondary aim is to quantify how vaccination and ventilation together reduce the pool of potential infectors in each of the settings by estimating the minimum viral emission rate needed to reproduce infection at varying levels of susceptibility.

## Materials and Methods

### Approach and Definitions

To develop our estimates, we defined a representative exposure scenario for each of the three settings (classroom, prison, restaurant) involving one infectious occupant in a room of typical geometry. We used an established airborne infection risk model to calculate the individual risk of infection (R) for each susceptible occupant, and the event reproduction number (R_event_) at varying ventilation rates and number of susceptibles. R_event_ is the expected number of new infections arising from a single infectious occupant at an event [4]. This is distinct from the basic reproduction number (R_0_), defined as the average number of new infections resulting from the introduction of a single infectious individual into a fully (100%) susceptible host population [5]. For modeling purposes, we quantified the number of susceptibles as the percent of the total occupants who are susceptible to infection (i.e., not successfully vaccinated or immune from prior infection). We use the term area concentration of susceptibles to represent the area of indoor space (square meters [m^2^]) per susceptible occupant. The threshold number of susceptibles and the threshold area concentration of susceptibles occur at a calculated R_event_ of one, above which one case begets more than another. For each setting we calculated these two threshold values at a mechanical ventilation rate based on American National Standards Institute (ANSI)/American Society of Heating, Refrigerating and Air-Conditioning Engineers (ASHRAE) 62.1 standards for acceptable air quality [6], and at a natural ventilation rate when windows cannot be opened and air exchange results solely from infiltration through the building envelope. We then determined the vaccination threshold as the complement of the threshold number of susceptibles assuming no immunity from prior infection. For comparative purposes, we also calculated the threshold values in each setting at a ventilation rate of 15 liters per second per person (L s^-1^ p^-1^), a typical goal for high indoor air quality consistent with EN 15251 Category I criteria for a non low polluting building [7].

### Calculation of the Event Reproduction Number (R_event_)

We used the Gammaitoni and Nucci [8] equation coupled with a Poisson dose-response model to calculate R_event_ for SARS-CoV-2 in a prototypical classroom, prison cell block, and full-service restaurant. The first step is calculating the probability of infection (P_I_) resulting from each exposure through equations (1-3):

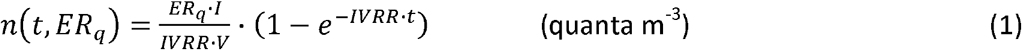

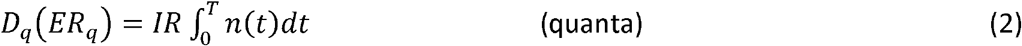

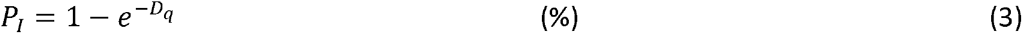

Where n represents the quanta (infectious dose for 63% of susceptible occupants by droplet nuclei inhalation) concentration in air at time t, ER_q_ is the quanta emission rate (quanta h^-1^), I is the number of infectious occupants (assumed to be only one), V is the volume of the indoor environment considered (m^3^), IVRR (h^-1^) represents the infectious virus removal rate in the space investigated, D_q_ is the dose of quanta inhaled by susceptible occupants, T is the total time of the exposure (h), and P_I_ is the probability of infection of a susceptible occupant. The infectious virus removal rate is the sum of the air exchange rate (AER) via ventilation in units of air changes per hour, the particle deposition on surfaces (k_d_, e.g. via gravitational settling), and the viral inactivation in ambient air (λ).

With all other parameters held constant, the probability of infection calculated in equation (3) assumes different values based on ER_q_. To evaluate the individual risk (R) of infection of an exposed susceptible occupant for a given exposure scenario, we then quantify the probability of infection as a function of ER_q_ (P_I_[ER_q_]) and the probability of occurrence of each ER_q_ value (P_ERq_) which can be defined by the probability density function (pdf_ERq_) of ER_q_ assuming a lognormal distribution. Since the probability of infection (P_I_[ER_q_]) and the probability of occurrence P_ERq_ are independent events, R for a given ER_q_, R(ER_q_), can be evaluated as the product of the two terms:

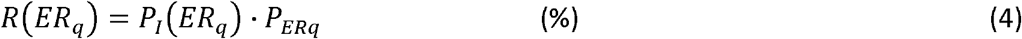

where P_I_(ER_q_) is the conditional probability of the infection, given a certain ER_q_, and P_ERq_ represents the relative frequency of the specific ER_q_ value. The individual risk (R) of an exposed susceptible occupant is then calculated by integrating the pdf_R_ for all possible ER_q_ values, i.e. summing up the R(ER_q_) values calculated in eq. (5):

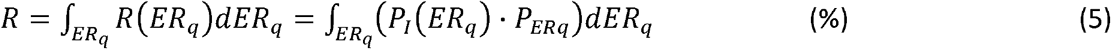

Equation (5) represents a numerical solution approximately equaling the average P_I_ that would result from a Monte Carlo simulation randomly sampling ER_q_ from its lognormal distribution. The individual risk R also represents the ratio between the number of new infections and the number of exposed susceptible occupants (S) for a given exposure scenario and considering all possible ER_q_ values from its lognormal distribution for the infectious occupant under investigation. For a single exposure event involving a single infectious occupant, R_event_ is calculated as the product of R and S as in eq. (6):

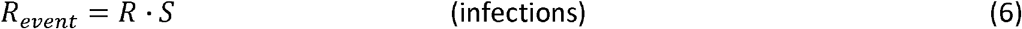

For a specific event, the threshold number of susceptibles occurs at the value of S where R_event_ equals one (S_threshold_ = 1/R) and is calculated by dividing S_threshold_ by the total room occupancy less the infected occupant. The threshold area concentration of susceptibles is calculated by dividing S_threshold_ by the room area.

### Modeling Scenario Input Parameters

Input parameters for the classroom, prison, and restaurant scenarios are summarized in Table 1. Geometry and default occupancy for the classroom are based on the rooms studied by Wells [1] with an exposure time of 5.5 hours representing a single day. The restaurant model encompasses the dining room geometry of the US Department of Energy building prototype for a full-service restaurant, with an exposure time of 1.5 hours [9]. The prison model is based on the largest cell block size studied by Hoge et al. [10], which was overcrowded with a median living area of 3.2 m^2^ per inmate. The exposure time for the prison scenario is likely highly variable, but we assume it to be 36 hours since inmates share the same airspace for extended time periods and peak infectiousness has been estimated to occur at 2 days before to 1 day after symptom onset [11]. Thus, a 36-hour period where infectiousness is at or near peak but without symptoms that would prompt quarantine can be reasonably expected.

**Table 1.** Modeling input and ventilation reference parameters

|  | Classroom | Prison | Restaurant | Average |
| --- | --- | --- | --- | --- |
| Room Volume ($\text{m}^3$) | 170 | 576 | 640 | 462 |
| Room Area ($\text{m}^2$) | 57 | 160 | 213 | 143 |
| Occupancy (Persons) | 20 | 50 | 100 | 57 |
| Occupancy ($\text{m}^2 \text{ Person}^{-1}$) | 2.8 | 3.2 | 2.1 | 2.7 |
| Exposure Time (h) | 5.5 | 36 | 1.5 | 14 |
| Infectious Occupant Activity | Standing, speaking | Resting, oral breathing | Resting, loudly speaking | -- |
| Median $\text{ER}_q \log_{10}$ (quanta $\text{h}^{-1}$) | 0.41 | -0.28 | 1.2 | 0.44 |
| Natural Ventilation AER ($\text{h}^{-1}$) | 0.5 | 0.5 | 0.5 | 0.5 |
| Mechanical Ventilation AER ($\text{h}^{-1}$) | 2.6 | 1.4 | 3.2 | 2.4 |
| High Air Quality AER ($\text{h}^{-1}$) | 6.4 | 4.7 | 8.4 | 6.5 |
| Natural Ventilation ($\text{L s}^{-1} \text{ p}^{-1}$) | 1.2 | 1.6 | 0.89 | 1.2 |
| Mechanical Ventilation ($\text{L s}^{-1} \text{ p}^{-1}$) | 6.1 | 4.4 | 5.7 | 5.4 |
| High Air Quality Ventilation ($\text{L s}^{-1} \text{ p}^{-1}$) | 15 | 15 | 15 | 15 |

The distributions for the quanta emission rate were modified from Buonanno et al. [12; see Supplemental Material] for standing and speaking for the classroom (assuming the class instructor is the emitting subject), resting and loudly speaking for the restaurant, and resting and oral breathing for the prison, with the log_10_ average ER_q_ values indicated in Table 1 and a log_10_ standard deviation for all distributions of 1.2. All susceptible occupants were assumed to be at rest with an inhalation rate of 0.49 m^3^ h^-1^. We used a deposition rate, k_d_, of 0.24 h^-1^ based on the ratio between the settling velocity of super-micrometer particles (roughly 1.0 × 10^−4^ m s^-1^ [13]) and the height of the emission source (1.5 m). For the SARS-CoV-2 inactivation rate in air, we used a value of 0.63 h^-1^ based on the measurements reported by van Doremalen et al. [14]. For each scenario, we varied the AER from zero to a maximum of six air changes per hour to calculate R and R_event_ at a number of susceptibles ranging from 0-100%.

## Results

The results of our modeling analysis are summarized in Figure 1 and Table 2 for each setting at an assumed natural ventilation rate of 0.5 air changes per hour, and at a mechanical ventilation rate corresponding to the applicable standard for acceptable air quality based on ANSI/ASHRAE 62.1 and shown in Table 1 [6]. The naturally ventilated restaurant (Figure 1A) has the lowest threshold number of susceptibles of 15%, and the mechanically ventilated classroom (Figure 1B) has the highest threshold number of susceptibles of 60%. The threshold number of susceptibles for the prison cell block (Figure 1C) exhibits the smallest difference between the natural ventilation (23%) and mechanical ventilation (31%) scenarios.

**Table 2.** Modeling results

|  | Ventilation | Classroom | Prison | Restaurant | Average |
| --- | --- | --- | --- | --- | --- |
| Individual Risk (R) (%) | Natural | 14% | 8.9% | 6.8% | 9.9% |
|  | Mechanical | 8.8% | 6.5% | 4.1% | 6.5% |
|  | High Air Quality | 5.5% | 3.4% | 2.3% | 3.7% |
| Threshold Number of Susceptibles (%) | Natural | 37% | 23% | 15% | 25% |
|  | Mechanical | 60% | 31% | 25% | 39% |
|  | High Air Quality | 95% | 60% | 44% | 66% |
| Threshold Area Concentration (m<sup>2</sup> Susceptible<sup>-1</sup>) | Natural | 8.1 | 14 | 14 | 12 |
|  | Mechanical | 5.0 | 11 | 8.6 | 8.2 |
|  | High Air Quality | 3.1 | 5.4 | 4.9 | 4.5 |

**Figure 1.**
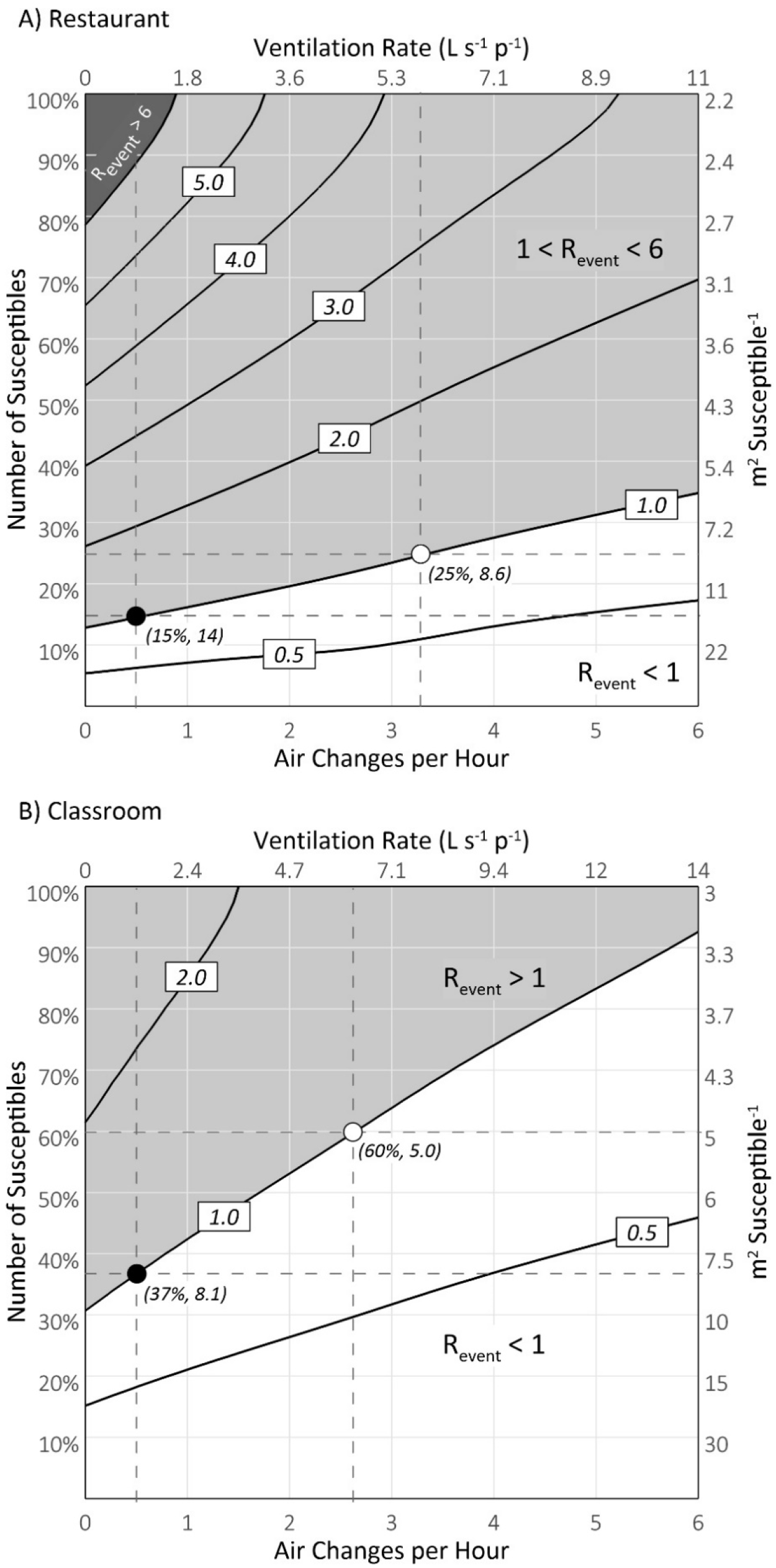
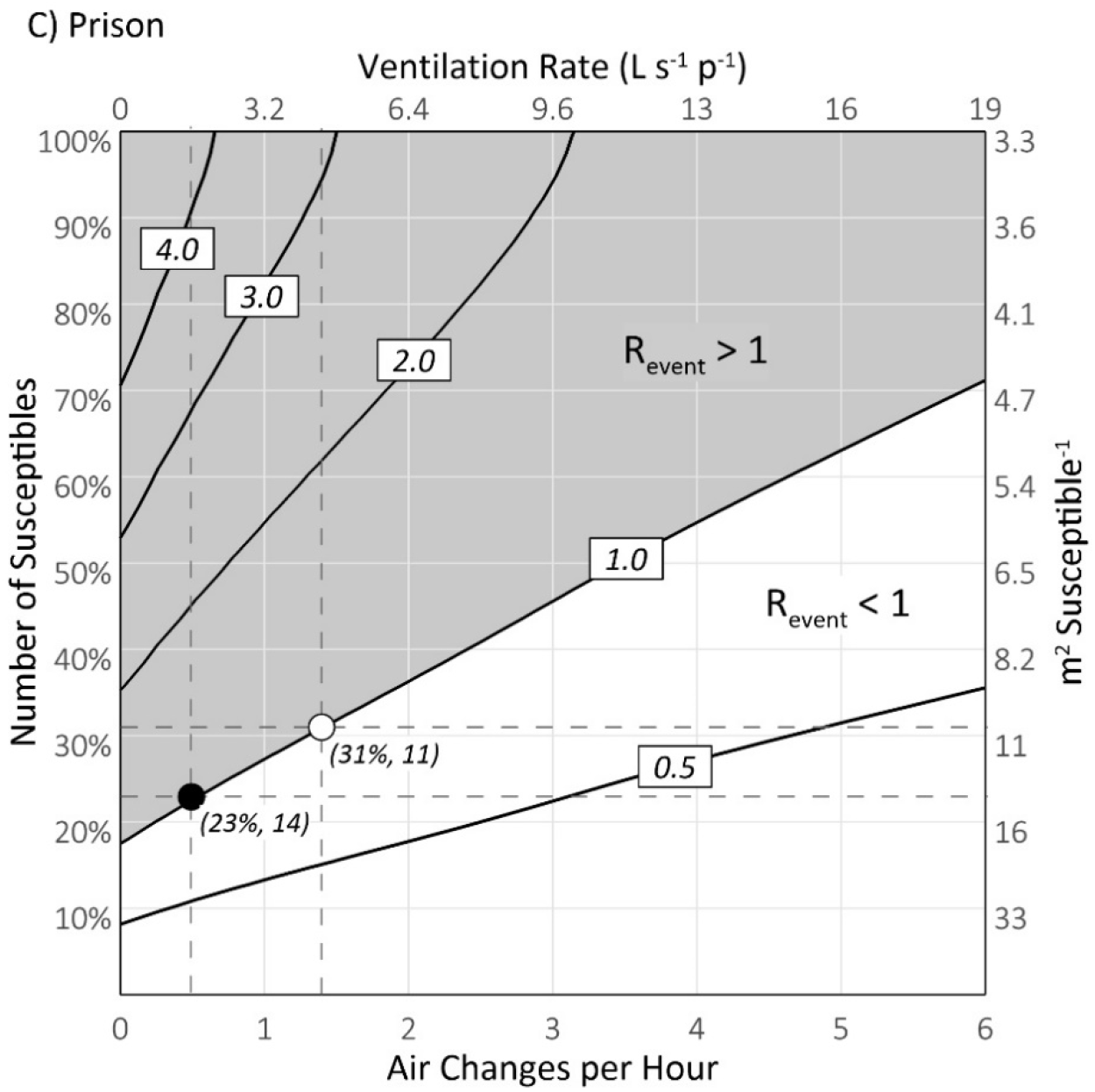
Surface graphs of R_event_ for SARS-CoV-2 as a function of the number of susceptibles and air exchange rate (AER) for the restaurant (A), classroom (B) and prison cell block (C) modeling scenarios. Contour lines connect equal R_event_ values. The black- and white-filled points along the R_event_ = 1.0 contour line identify the threshold number of susceptibles for natural ventilation and mechanical ventilation scenarios, respectively, at the intersection of the dashed horizontal and vertical lines. The threshold values are labeled in parenthesis in terms of both the percent susceptible and m^2^ susceptible^-1^.

The average threshold number of susceptibles for all three settings calculated under the natural and mechanical ventilation rates is 32%. In the absence of immunity from prior infections and assuming vaccination confers complete protection, these results suggest an average vaccination threshold of 68% with a range of 40-85%. The naturally ventilated prison and restaurant have the highest threshold area concentration of susceptibles at 14 m^2^ susceptible^-1^, while the mechanically ventilated classroom has the lowest at approximately 5.0 m^2^ susceptible^-1^. The overall average threshold area concentration of susceptibles for mechanical and natural ventilation is approximately 10 m^2^ susceptible^-1^.

Increasing the ventilation rate to the high air quality metric of 15 L s^-1^ p^-1^ increases the threshold number of susceptibles to 95% in the classroom, 60% in the prison, and 44% in the restaurant. The average threshold number of susceptibles for all three settings becomes 66%, more than twice the average of the natural and mechanical ventilation scenarios. To maintain an R_event_ of one in a fully susceptible population, the estimated ventilation requirements are 43 L s^-1^ p^-1^ (24 air changes per hour), 30 L s^-1^ p^-1^ (9.5 air changes per hour) and 17 L s^-1^ p^-1^ (7.0 air changes per hour) for the restaurant, prison, and classroom, respectively. Such high air exchange rates are impracticable in many settings, suggesting a role for ultraviolet air disinfection [15, 16].

Increasing ventilation and/or decreasing the number of susceptibles has the effect of increasing the minimum ER_q_ necessary to produce an R_event_ of one, thereby reducing the number of infected occupants capable of infecting others on average. This is illustrated in Figure 2 for the prison cell block model. For the naturally ventilated cell block in a fully susceptible population, the minimum ER_q_ is just below 1.0 quanta h^-1^, occurring at the 58^th^ percentile value of the resting, oral breathing distribution. At a number of susceptibles of 23%, the minimum ER_q_ becomes approximately 4.3 quanta h^-1^ at the 78^th^ percentile value. Increasing ventilation to 15 L s^-1^ p^-1^ further decreases the pool of potential infectors, raising the minimum ER_q_ to approximately 17 quanta h^-1^ at the 90^th^ percentile value, indicating only a 10% chance of a secondary infection.

**Figure 2.**
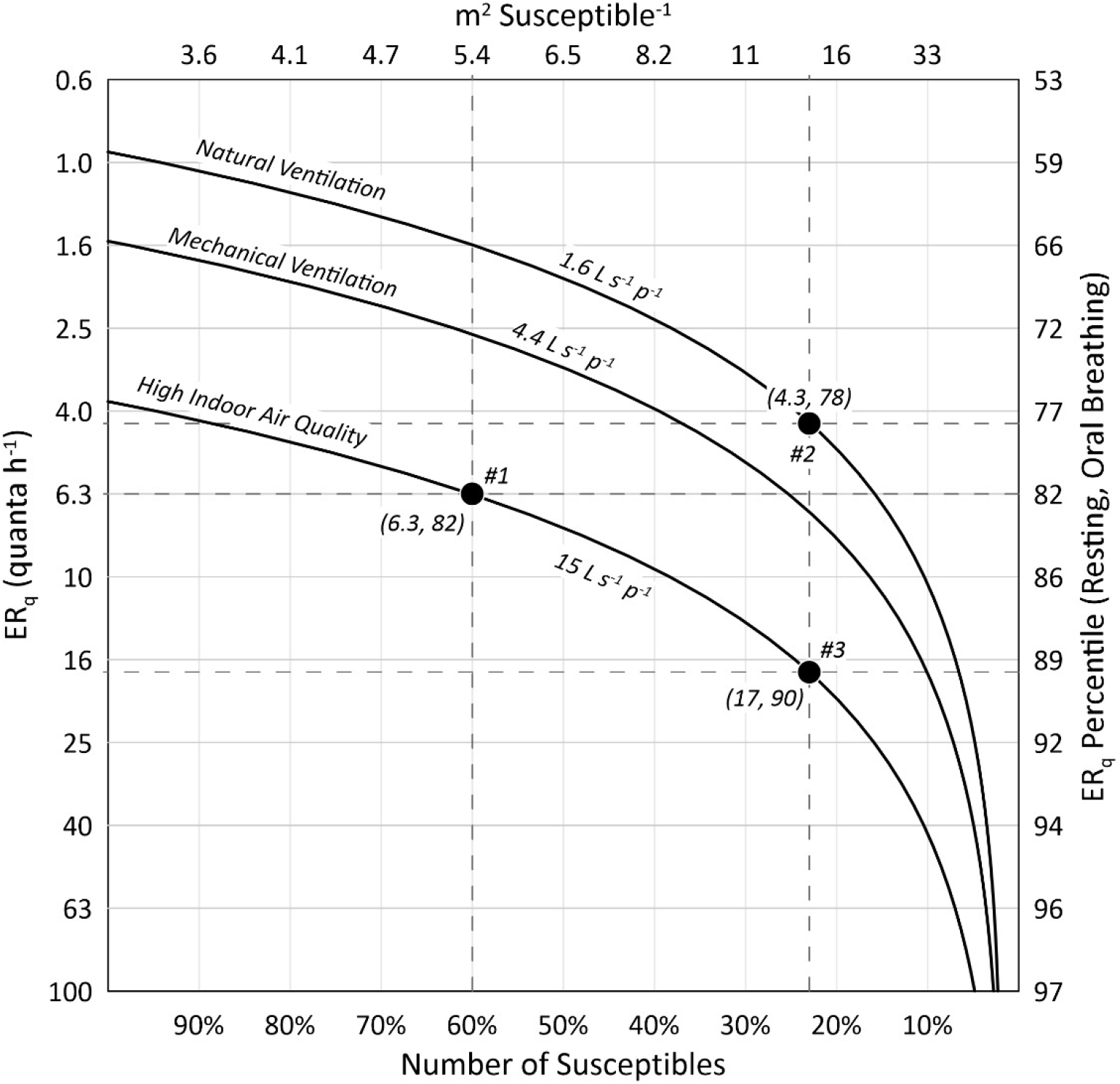
Minimum quanta emission rates (ER_q_) for R_event_ ≥ 1.0 for the prison scenario under natural ventilation, mechanical ventilation, and high air quality ventilation conditions as a function of the number of susceptibles. Points #1 and #2 identify the minimum emission rates for high air quality ventilation and natural ventilation at their respective threshold number of susceptibles from Figure 1C. Point #3 identifies the minimum emission rate for high air quality ventilation at the natural ventilation threshold number of susceptibles, representing both high ventilation and high vaccination. The minimum emission values are labeled in parenthesis, denoting the emission in quanta h^-1^ and its corresponding percentile in the resting, oral breathing ER_q_ distribution.

## Discussion

The overall average threshold number of susceptibles calculated for the natural and mechanical ventilation scenarios is 32%, suggesting a basic reproduction number (R_0_) of approximately 3 in accordance with general epidemiological theory that the equilibrium susceptible fraction in a host population is the reciprocal of R_0_ [5]. This is consistent with R_0_ estimates for the initial SARS-CoV-2 outbreaks in Wuhan, China and Northern Italy [17]. Our analysis is also consistent with the overdispersed epidemiological nature of SARS-CoV-2 [18], with a minority of cases accounting for most secondary transmissions. In the naturally ventilated prison, we calculate that emissions approximately below the 60^th^ percentile value will fail to reproduce infection, on average, indicating the median emission is not a significant source of transmission (Figure 2). Furthermore, application of equation (5) for the prison scenario shows that emissions above the 80^th^ percentile value account for at least 85% of the total individual risk, suggesting a dispersion parameter (k) between 0.10 and 0.16. This derivation is provided in the Supplemental Material and enables quantification of the probability of SARS-CoV-2 superspreading and outbreak extinction as defined by Lloyd-Smith et al. [19]. Due to this overdispersion, vaccinating 77% of inmates in a naturally ventilated cell block still leaves the remaining susceptible population vulnerable to emitters above the 78^th^ percentile. As a result, explosive but comparatively rare superspreading events may continue in crowded, poorly ventilated settings, a phenomenon that challenges the eradication of measles virus [20].

Applying both high vaccination and high ventilation raises both the threshold number of susceptibles and the minimum emission rate needed to reproduce infection, decreasing the dispersion parameter and increasing the probability of outbreak extinction. Uniformly increasing ventilation to a high air indoor air quality metric of 15 L s^-1^ p^-1^ approximately doubles the average threshold number of susceptibles and therefore halves vaccination requirements for equivalent prevention of infection. Thus, while a ventilation rate of 15 L s^-1^ p^-1^ is unlikely to prevent all secondary infections when a high-emitting index case is introduced into a fully susceptible, indoor population [21], it can provide a substantial downstream epidemiological benefit relative to a poorly ventilated baseline condition. This effect is important for pathogens where transmission is overdispersed, with *Mycobacterium tuberculosis* being another example [22], as superspreading events (SSEs) facilitate infection of the high-emitting minority that continues the chain of contagion. For our prison cell block model, we estimate that increasing the natural ventilation rate to the high air quality ventilation rate decreases the SSE probability from 16% to 6.6% (see Supplemental Material). This is an important finding, as prisons and jails are clear hot spots for SARS-CoV-2 transmission. For example, by March 2021, five California State Prisons (Chuckawalla Valley, California Rehabilitation Center, Avenal, San Quentin, and California Men’s Colony) reported total confirmed COVID-19 case rates above 800 per 1,000 inmates [23]. Such high case rates imply a low threshold number of susceptibles, with inadequate ventilation a likely factor. Indeed, during an investigation of the San Quentin State Prison in June 2020, McCoy et al. [24] noted cell blocks with windows that were welded shut and with fan systems that appeared to have been inactive for years.

Historical examples for measles virus illustrating the relationship between ventilation and the threshold number of susceptibles in classrooms are provided by Wells [1, 25] and Thomas [26]. In classic experiments using upper-room air irradiation in primary and upper school classrooms during the 1941 outbreak of measles in suburban Philadelphia, USA, Wells estimated a threshold number of susceptibles of approximately 20% in unirradiated rooms at a then-standard ventilation rate of 14 L s^-1^ p^-1^. Irradiated classrooms supported a much higher threshold number of susceptibles of approximately 57% because the weekly probability of infection in the irradiated rooms was approximately four to five times lower than in the unirradiated rooms [1, 25]. The findings of Wells are similar to those of Thomas [26] who studied the spread of measles in primary schools in the Woolwich district of London in 1904. Thomas concluded that outbreaks of measles tend to occur when the number of susceptibles exceeds approximately 33% and generally continue until the proportion is reduced to 18%. However, the spread of measles in the Woolwich classrooms below the 33% threshold was highly heterogeneous, with many experiencing significant outbreaks infecting a majority of susceptible occupants. The three classes with a number of susceptibles below 10% experienced zero cases of measles, and two temporary schools with crowding and poor ventilation had explosive outbreaks that nearly exhausted the population of susceptibles, with a median probability of infection of 87% for the five classes in the two schools. Thomas measured a carbon dioxide (CO_2_) concentration of 3,000 parts per million in one of the temporary schools [26], indicating a steady-state ventilation rate below 2 L s^-1^ p^-1^ and comparable to our natural ventilation scenario. The higher contagiousness of measles as compared to SARS-CoV-2 is illustrated by the historical reported threshold number of susceptibles of 20-33% as compared to our classroom estimate of 37-60% despite the lower ventilation standards of present day. This difference is also reflected by the median classroom measles probability of infection of 87% for the poorly ventilated temporary schools studied by Thomas [26] as compared to the individual risk (R) of approximately 14% we calculated for SARS-CoV-2 (Table 2). A ventilation rate of 14 L s^-1^ p^-1^ appears sufficient, on average, to prevent sustained airborne transmission of SARS-CoV-2 in a classroom with a number of susceptibles up to approximately 90%.

A limitation of our infection risk modeling approach is the assumption of a homogeneous concentration of droplet nuclei within the room, with viral emissions being instantaneously and completely mixed. However, a recent comparison of this box-modeling approach with computational fluid dynamics (CFD) simulations for a classroom environment indicates relatively minor errors for natural (6%) and forced mechanical (29%) ventilation scenarios [27]. The uncertainty in the emission rate, based on viral loads that vary several orders of magnitude between individuals and over time [28], is likely much more significant than that caused by incomplete mixing at the small scale of our models. Further improvements to the emission rate distributions are needed that incorporate variation in droplet volume concentrations [28, 29], such that a more complete stochastic emission model can be implemented. An additional limitation is our estimation of vaccination thresholds using singular, setting-specific events, without considering cumulative exposure effects that may result from an infectious person attending class in two successive days, for example. The importance of singular SSEs on SARS-CoV-2 transmission is well established, and such events likely occur during a narrow 1-2 day window of peak infectivity [30]. As such we do not expect cumulative exposures to be a significant factor outside of co-habitation environments, which is why our prison scenario used a 36-hour duration. Our approach also does not account for extreme examples such as someone visiting multiple similar restaurants for similar durations on the same evening (thus increasing the number of exposed susceptibles to a similar infectious dose), or for a bartender or other vocalizing restaurant employee who may be present for much longer than 1.5 hours. Indeed, there are numerous other scenarios, such as choirs or high-intensity exercise rooms, where higher vaccination thresholds are likely, reinforcing the need for high levels of both vaccination and ventilation also considering that vaccines are not 100% protective.

## Conclusions

Our fully prospective airborne infection modeling results are consistent with the transmission dynamics of SARS-CoV-2 and illustrate the challenges presented by substantial heterogeneity in the settings of contagion and a skewed viral emission rate distribution. To support pre-pandemic levels of occupancy, required vaccination rates are much higher for a naturally ventilated restaurant (85%) than for a mechanically ventilated classroom (40%). As vaccination campaigns progress it follows that occupancy limitations should be relaxed for classrooms before full-service indoor restaurants. Maintaining focus on enhanced ventilation together with vaccination is especially important considering the emergence of new SARS-CoV-2 strains that are more contagious with increasing possibility of second infections or vaccine breakthrough infections. Avoidance of overcrowding remains a critical strategy to minimize airborne transmission, as our calculations suggest ensuring an average of 10 m^2^ per susceptible occupant of an indoor space is approximately equivalent to achieving a number of susceptibles of 32% of normal occupancy. This is because the ventilation rate per susceptible occupant is more than tripled relative to the baseline average occupant loading of 2.7 m^2^ per susceptible occupant for the three settings evaluated herein.

## Data Availability

The authors do not have permission to share the data.

## Declarations

### Ethics approval and consent to participate

Not applicable

### Consent for publication

Not applicable

### Availability of data and materials

All data generated or analysed during this study are included in this published article and its supplementary information file.

### Competing interests

The authors have no conflicts or competing interests to disclose. Funding: This manuscript was prepared without external funding.

### Authors’ contributions

AM, LS, GB, LM designed research; AM performed research; AM, LS, GB, LM analyzed data; AM wrote the paper; and LS, GB, LM reviewed the paper

## Acknowledgements

The authors thanks Chantal Labbé at QUT ILAQH for her invaluable research support.

## Notes

### Competing Interest Statement

The authors have declared no competing interest.

### Funding Statement

No funding.

### Author Declarations

No ethics was required.

