## Supplemental Material for "The Vaccination Threshold for SARS-CoV-2 Depends on the Indoor Setting and Room Ventilation"

### 1.0 Quanta Emission Rate Distributions

For the infection risk modeling performed in this paper we modified the quanta emission rate (ER_q_) distributions for SARS-CoV-2 for three different expiratory activities (resting, oral breathing; standing, speaking; and resting, loudly speaking) from the original estimates presented in previous works [1, 2]. Specifically, we updated the viral load in sputum (c_v_ [RNA copies mL^-1^]) and the conversion factor (c*_i_* [quanta RNA copies^-1^]) defining the ratio between one quantum and the infectious dose expressed in viral RNA copies. A quantum is equivalent to a human infectious dose by inhalation for 63% of susceptibles (HID_63_). For the viral load, we incorporated data from multiple studies [3-6] to calculate an average value of 5.6 log_10_ RNA copies mL^-1^. The standard deviations of the viral load distributions were not explicitly reported in the studies; hence, we calculated the standard deviation such that the 99.9^th^ percentile value of a lognormal distribution (or roughly three standard deviations above the mean) approximately equaled the average of the maximums reported in the studies, or 9.4 log_10_ RNA copies mL^-1^. This resulted in a log_10_ standard deviation of 1.2 log_10_ RNA copies mL^-1^. For the updated conversion factor, we used a value of 1.4 x 10^-3^ quanta RNA copies^-1^ based on the thermodynamic equilibrium dose-response model developed by Gale [7]. The values for the inhalation rate (IR) and droplet volume concentration (V_d_) remained unchanged from [2], with the product of these two terms (V_d_∙IR) equaling a droplet volume emission rate for the three expiratory activities as follows: 9.8 x 10^-4^ mL h^-1^ for resting, oral breathing; 4.9 x 10^-3^ mL h^-1^ for standing, speaking; and 2.9 x 10^-2^ mL h^-1^ for resting, loudly speaking.

The average log_10_ ER_q_ value for each activity was then calculated through equation (1) using the average c_v_ of 5.6 log_10_ RNA copies mL^-1^:

${ER}_{q}=c_{v}\cdot c_{i}\cdot IR\cdot V_{d}$	(quanta h^-1^)	(1)

As variation in the parameterization was limited to the viral load (c_v_), the log_10_ standard deviation for ER_q_ equals that of the viral load for all activities, or 1.2 log_10_ quanta h^-1^.

### 2.0 Evaluation of Risk Proportion and the Dispersion Parameter

Of epidemiological interest for population-level modeling of SARS-CoV-2 is the proportion of transmission caused by the most infectious 20% of cases and how it compares to the so-called ‘20/80 rule’ under which 20% of cases cause 80% of transmission [8, 9]. In our airborne contagion model, the use of a defined lognormal distribution of the viral emission rate (ER_q_) allows quantification of the proportion of the individual risk (R) caused by the highest 20% of emissions (i.e., from ER_q_ values above the 80^th^ percentile). With the emission rate as a surrogate for individual infectiousness and assuming airborne transmission is dominant, we can thus compare the proportion of population-level transmission to the proportion of individual risk in a specific setting. Our model of a prison cell block is best suited for this analysis because it can be approximately considered a closed community, and an infectious occupant likely spends the majority of his or her peak infectious period in the same atmosphere as the susceptible occupants in the community. Supporting this assumption, our calculated value of R_event_ for the mechanically ventilated cell block in a fully susceptible population is 3.2, consistent with common estimates of R_0_ for SARS-CoV-2.

Supplementary Figure S1 presents the proportion of the individual risk as a function of the proportion of the ER_q_ distribution for resting, oral breathing for the prison cell block scenario. The ER_q_ proportion is ranked by infectiousness, such that a proportion of 0.2 represents the contribution from emissions above the 80^th^ percentile. The model results for the probability of infection as a function of ER_q_ are also presented for reference. The proportion of risk caused by the highest 20% of emissions (r_20_) ranges from a low of 85% for the natural ventilation scenario to a high of 90% for the high air quality scenario, with an average of 87% for the three scenarios. The risk proportion curve for the mechanical ventilation scenario closely resembles the expected proportion of transmission curve generated by Lloyd-Smith et al. [9] for SARS-CoV-1, indicating that the data can be reasonably modeled with a negative binomial distribution. This result is not unexpected, as the probability of infection is calculated using a Poisson process with a skewed ER_q_ distribution generating the primary input parameter.

**Supplementary Figure S1**

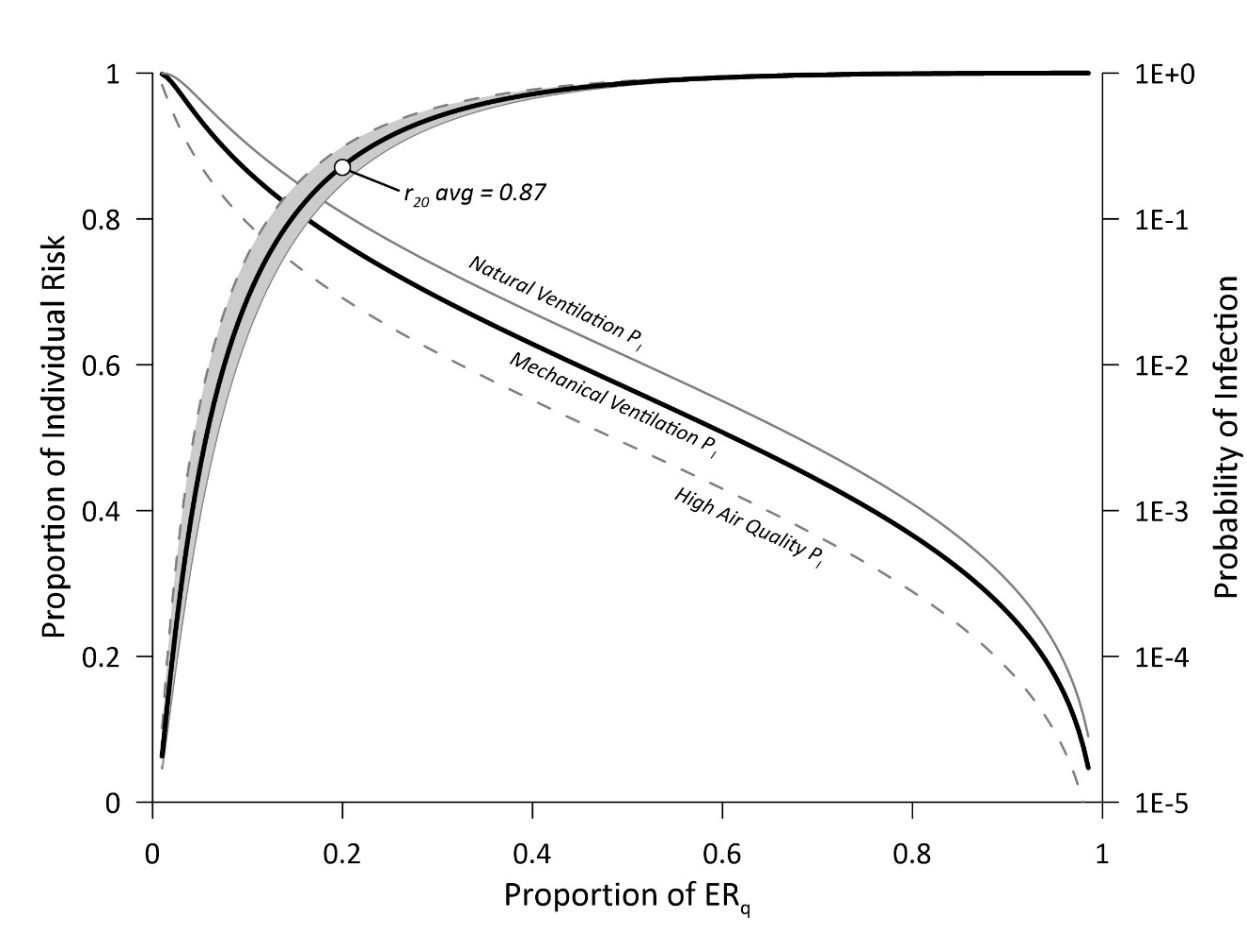

**Supplementary Figure S1** **Caption** – Proportion of the individual risk (R) contributed by the highest 20% of the ER_q_ distribution (r_20_), and the probability of infection (P_[I]_) for the prison cell block scenario. Solid black line represents mechanical ventilation, solid gray line represents natural ventilation, and dashed line represents natural ventilation. The proportion of the individual risk curves are filled with gray shading. Note the log_10_ scale on the secondary Y-axis corresponding to the probability of infection curves.

To estimate the negative binomial distribution dispersion parameter, k, from the data, we employed the following equation (2) for the probability generating function (g[s]) of the secondary infection (offspring) distribution from Lloyd-Smith et al. [9]:

$g(s)=\left( 1+\frac{R_{event}}{k}\left( 1-s \right) \right)^{-k}$	(%)	(2)

Using the R_event_ results from our modeling scenarios we solved for k for the special case of g(0), which equals the probability the infectious occupant fails to infect anyone else, or the proportion of zeroes. In our case, g(0) is assumed to equal the percentile value from the ER_q_ distribution corresponding to R_event_ = 1 in a fully susceptible population (i.e., the minimum ER_q_ necessary to produce one secondary infection on average, obtained from Figure 2 of the main text). The results for the dispersion parameter fitting analysis are presented on Supplementary Figure 2, with the y-intercept of each line equaling the probability of zero secondary infections resulting from the introduction of an infectious occupant into the prison cell block. The range of estimated k values is 0.10 – 0.16, consistent with estimates for both SARS-CoV-1 [9] and SARS-CoV-2 [10] and demonstrating the overdispersed nature of SARS-CoV-2 with a minority of individuals causing most secondary cases.

Also noted on Supplementary Figure S2 is the point on each curve where g(s) = s, which according to the negative binomial branching process signifies the probability of stochastic outbreak extinction, q [9]. The extinction risk, q, increases significantly with increasing ventilation, with the high air quality ventilation rate of 15 L s^-1^ p^-1^ producing an extinction risk of 91% as compared to 70% for the naturally ventilated cell block.

**Supplementary Figure S2**

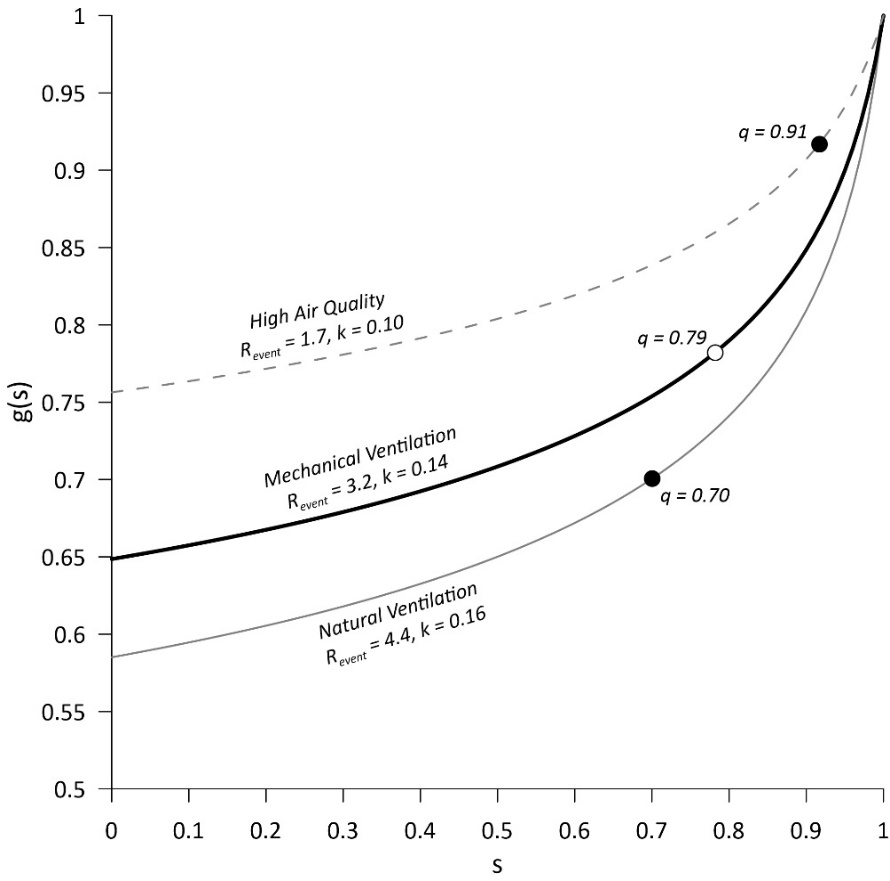

**Supplementary Figure S2** **Caption** – The probability generating functions of offspring distributions as defined by Lloyd-Smith et al. [9] plotted for the R_event_ values for the three different ventilation rates and a dispersion parameter, k, that sets the y-intercept equal to the minimum percentile value of the ER_q_ distribution resulting in R_event_ ≥ 1. Values of the outbreak extinction probability, q, are labeled at the point on each curve where g(s) = s [9].

Supplementary Figure S3 presents the secondary transmission probabilities for the three ventilation rates for the prison scenario, based on the percentiles of the lognormal ER_q_ distribution, exhibiting the characteristic negative binomial shape. Of interest is the probability of exceeding 8 secondary cases, which represents a superspreading event (SSE) threshold using the definition that an SSE causes more infections than the 99^th^ percentile of a Poisson distribution for an R_0_ of approximately 3.0 [9, 11]. Increasing the natural ventilation rate to the high air quality ventilation rate decreases the SSE probability from 16% to 6.6%.

**Supplementary Figure S3**

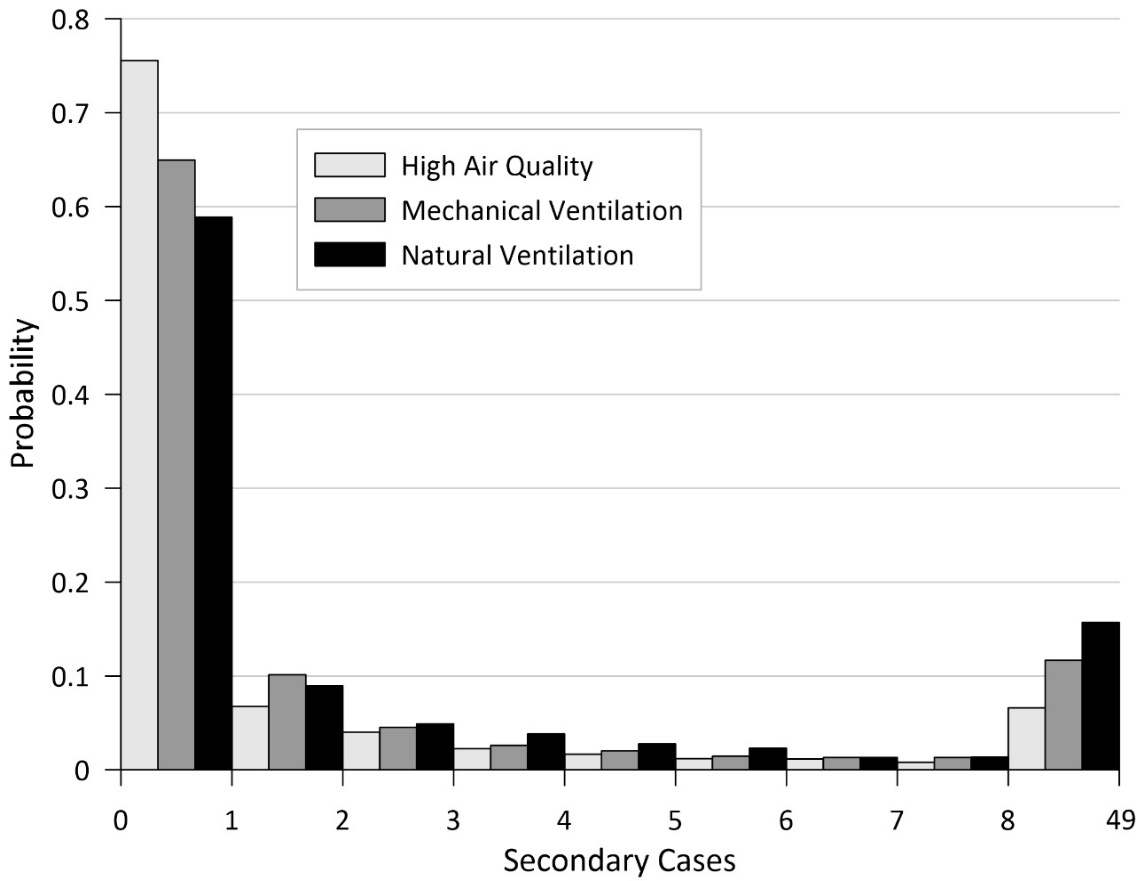

**Supplementary Figure S3 Caption** – Secondary transmission probabilities for the prison scenario with 49 susceptible occupants (100% number of susceptibles). The probability of exceeding 8 secondary cases represents the superspreading event (SSE) probability.

Supplementary Table S1 summarizes the statistical results of our analysis of the introduction of a single infectious occupant in a prison cell block with 49 susceptible occupants at varying levels of ventilation. We note that these estimates apply to a fully susceptible population, and with the addition of vaccination and immunity from prior infection, secondary transmission and SSE probabilities decrease. To illustrate this, Supplementary Figure S4 presents the secondary transmission probabilities for the prison scenario at the natural ventilation threshold number of susceptibles of 23%. Combining the highest evaluated levels of both ventilation and vaccination reduces the secondary transmission probability to approximately 10% (as also shown on Figure 2 of the main text) and reduces the SSE probability to approximately 1.5%.

**Supplementary Table S1** – Summary of airborne infection risk calculations for the introduction of a single infectious occupant into an otherwise fully susceptible prison cell block.

| Ventilation | Individual Risk | R_event_ | Probability of Zero Secondary Cases | Probability of Superspreading Event | Probability of Outbreak Extinction |
| --- | --- | --- | --- | --- | --- |
| Natural | 8.9% | 4.4 | 58% | 16% | 70% |
| Mechanical | 6.5% | 3.2 | 65% | 12% | 79% |
| High Air Quality | 3.4% | 1.7 | 76% | 6.6% | 91% |
| Average | 6.3% | 3.1 | 66% | 12% | 80% |

**Supplementary Figure S4**

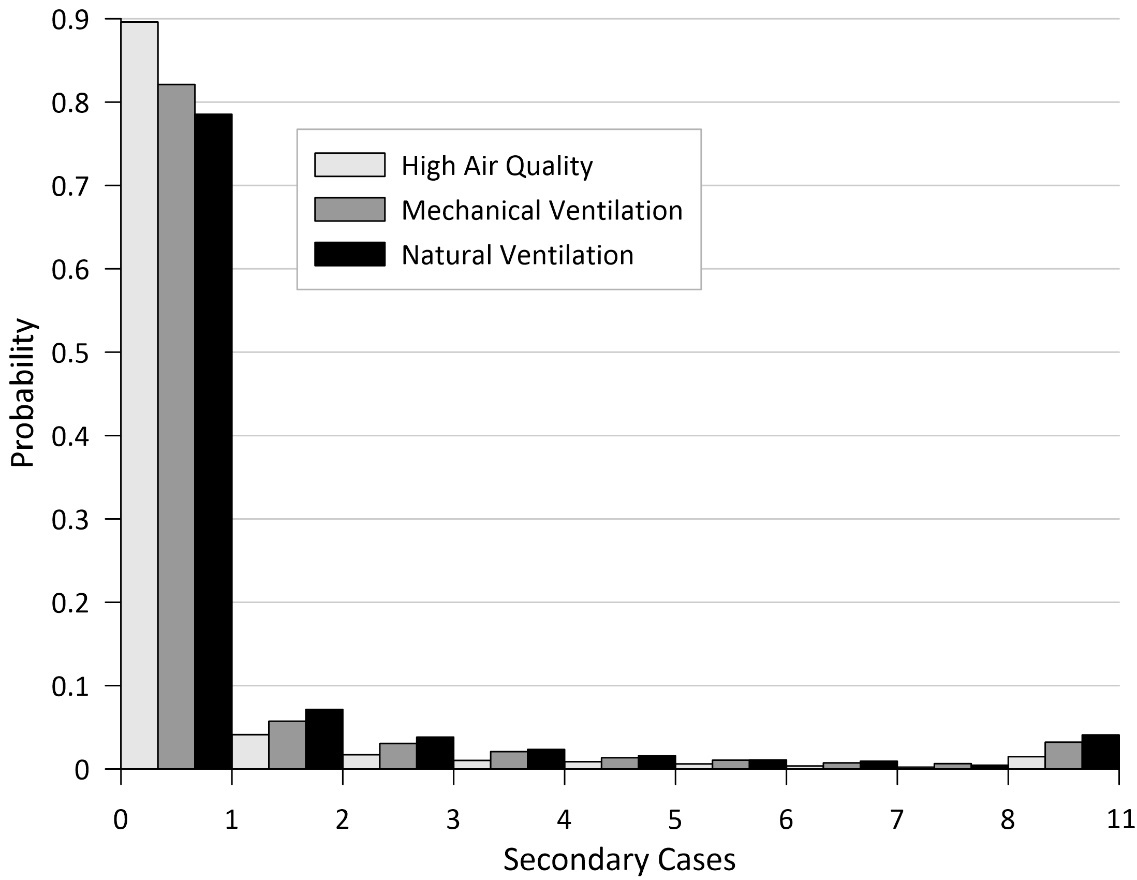

**Supplementary Figure S4 Caption** – Secondary transmission probabilities for the prison scenario with 11 susceptible occupants (23% number of susceptibles).
